# Development of a core measurement set for research in degenerative cervical myelopathy: a study protocol (AO Spine RECODE-DCM CMS)

**DOI:** 10.1101/2021.11.11.21266170

**Authors:** Benjamin M. Davies, Alvaro Yanez Touzet, Oliver D. Mowforth, Keng Siang Lee, Danyal Khan, Julio C. Furlan, Michael G. Fehlings, James Harrop, Carl M. Zipser, Ricardo Rodrigues-Pinto, James Milligan, Ellen Sarewitz, Armin Curt, Vafa Rahimi-Movaghar, Bizhan Aarabi, Timothy F. Boerger, Lindsay Tetreault, Robert Chen, James D. Guest, Sukhvinder Kalsi-Ryan, Iwan Sadler, Shirley Widdop, Angus G. K. McNair, Mark R. N. Kotter, On behalf of the AO Spine RECODE-DCM Steering Committee

## Abstract

**Introduction:** Progress in degenerative cervical myelopathy (DCM) is hindered by inconsistent measurement and reporting of outcomes. This can, for example, impede the aggregation of data and comparison of outcomes between studies. This limitation can be reversed by developing a core measurement set (CMS) for use in DCM research. Previously, the AO Spine Research Objectives and Common Data Elements for DCM (AO Spine RECODE-DCM) defined ‘what’ should be measured in DCM: specifically, the core data elements and core outcome set of the disease. The next step of this initiative is to determine ‘how’ to measure these features. The current protocol outlines the steps necessary for the development of a CMS for DCM research and audit.

**Methods and analysis:** The CMS will be developed in accordance with the guidance developed by the Core Outcome Measures in Effectiveness Trials (COMET) and the Consensus-based Standards for the selection of health Measurement Instruments (COSMIN). The process will involve five phases: (1) agreement on the measurement constructs and approaches to their evaluation; (2) the formation of a long list of potential measurement instruments, by identifying existing instruments and assessing their psychometric properties; (3) the aggregation of evidence concerning ‘when’ measurements should be taken; (4) consensus about which instruments to include in the CMS; and (5) implementation.

**Ethics and dissemination:** Ethical approval was obtained from the University of Cambridge. Dissemination strategies to promote awareness and adoption of the CMS include peer-reviewed scientific publications; conference presentations; podcasts; the identification of AO Spine RECODE-DCM ambassadors; and engagement with relevant journals, funders, and the DCM community.

**Impact of this work:** The proposed project will enable standardised and comprehensive measurement in DCM clinical trials. The CMS will be established using a robust, global, and multi-stakeholder consensus process, with broad representation of healthcare professionals and individuals living with the disease. It will focus on measurement instruments currently in use. This ensures that the CMS will be immediately usable and suited for widespread adoption. The development of better outcome instruments in DCM remains a top 10 research priority and this work will hence facilitate knowledge generation for this important disease.

## INTRODUCTION

### Background

Degenerative cervical myelopathy (DCM) is a common and often disabling disease (1). Estimated to affect as many as one in fifty adults (1), it develops due to degenerative and/or congenital changes in the cervical spine leading to mechanical stress and a progressive spinal cord injury (2-4). This disease can lead to a wide variety of symptoms, affecting the whole body (5). These symptoms commonly include gait dysfunction, imbalance and falls, loss of strength and manual dexterity, and pain. Despite current best practice (6), a minority of patients will make a full recovery and DCM is often associated with lifelong disability, impaired quality of life, and significant costs to both the individual and to society (7, 8).

Whilst progress has been and is being made (6, 9), there remain significant knowledge gaps. For people affected by DCM, solutions to these challenges cannot come soon enough (10). AO Spine Research Objectives and Common Data Elements for Degenerative Cervical Myelopathy (AO Spine RECODE-DCM; www.aospine.org/recode) is an international, multi-stakeholder initiative originally formed to create a ‘research toolkit’ that could help accelerate knowledge discovery and improve outcomes in DCM (11, 12). This project aimed to unify terminology, outcome measurement, and reporting (12-14) in order to enable data aggregation and implementation of management recommendations (15-17). The value of addressing these inefficiencies is likely magnified for DCM, as the research community is relatively small, fragmented, and has not received commensurate attention or funding (18, 19).

So far, AO Spine RECODE-DCM has established the top research priorities and agreed on a single definition and index term. It has also agreed on ‘what’ should be measured in DCM research: that is, a minimum data set, which is comprised of core data elements (CDE) and a core outcome set (COS). The COS is composed of six domains: neuromuscular function, life impact, pain, radiology, economic impact and adverse events. Each domain contains a list of more specific outcomes that should be measured. Whilst adherence to this minimum dataset should ensure a more comprehensive assessment of DCM, to ensure data is reported in a consistent manner, best suited for between study comparison and evidence synthesis, this standardisation should also extend to ‘how’ the dataset should be measured and reported. This additional phase is referred to as the development of a core measurement set (CMS) (20-22).

A CMS is a set of agreed upon tools that are used to measure the CDE and COS (23). A CMS is needed to improve the consistency of data measurement and reporting across DCM and will ultimately accelerate changes that will improve outcomes for this population (12). This protocol defines how AO Spine RECODE-DCM will establish a CMS for DCM.

## METHODS AND ANALYSIS

### Overview and scope

The CMS will continue to be managed within the framework of AO Spine RECODE-DCM (11). Ethical approval for this project was obtained from the University of Cambridge (Ethical approval number: HBREC2019.14). A multi-disciplinary, global steering committee (SC) was formed for the oversight of the project (www.aospine.org/recode). In addition to interim correspondence, the committee meets at least twice a year. For a meeting to be considered quorate, it must include at least two people with lived experience and four healthcare professionals. When a steering group member is unable to attend, decisions made at quorate meetings are respected. Day-to-day administration is provided by a multi-stakeholder management group.

As outlined earlier, the standardisation of data measurement and reporting is an immediate priority for DCM. However, the research priority-setting process further recognised a need to develop new measurement instruments for DCM (24). Acknowledging that such development demands a significant period of time and financial support, it was decided that the initial CMS should focus on selecting the most relevant—but existing—instruments, as opposed to developing new tools or selecting those early in development. The added benefit would be to enable comparisons with historic data while simplifying the implementation of DCM’s first minimum dataset. This rationale is expanded in the discussion.

The development of the CMS is based on relevant guidance, including that developed by the Core Outcome Measures in Effectiveness Trials (COMET) and the Consensus-based Standards for the selection of health Measurement Instruments (COSMIN) (23, 25-32). Notably, no more than one measurement tool will be selected per core outcome (23). The developmental process will be conducted in five phases (**Figure 1**):

**Figure 1.**
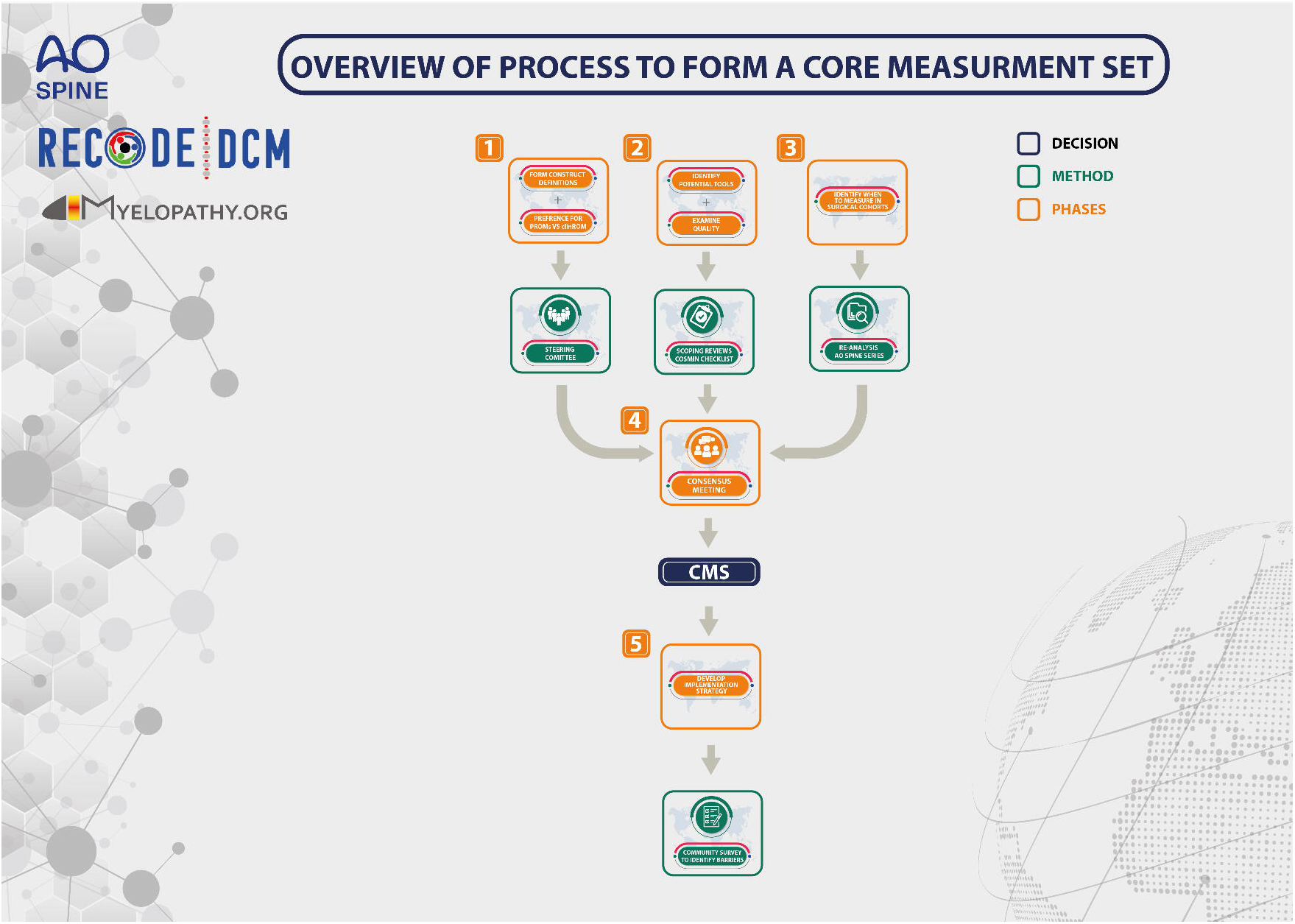
Overview of the CMS process. (1) Phase 1: To agree on the measurement construct and preferred measurement approach. (2) Phase 2: To identify measurement tools and evaluate their evidence base. (3) Phase 3: To aggregate the evidence on timing of assessment. (4) Phase 4: To select the most appropriate instruments through multi-stakeholder consensus and provide reporting guidance. (5) Phase 5: To implement the CMS

The CMS will cover each element contained within the CDE and COS (the minimum dataset). Elements in the CDE which are descriptive (e.g., individual’s age or sex) and do not require measurement *per se*, will only feature in Phases 3 and 4. These elements will be identified and agreed during Phase 1.

### Phase 1. Forming measurement constructs and establishing the preferred measurement approach

During the formation of the CDE and COS, each element was summarised with a lay description. Whilst this provided an explanation as to how the term was originally proposed, for example based on content from interviews (5, 10), these descriptions were not intended as construct definitions. Further, as some outcomes were merged and/or renamed during the process, they lacked a unifying explanatory statement.

Consequently, the first step of this CMS is to agree on the specific construct to be measured (23, 25-32). These will be expressed by forming a definition for each element. Draft definitions will be generated from original source documents including published literature or interviews with patients and professionals. This will be undertaken by the management group. These provisional definitions will then be reviewed by the SC and iterated as indicated. Each definition must reach >70% approval at a quorate meeting to be considered final.

For elements requiring measurement, the SC will also define through agreement, whether it should be ideally measured by people with DCM (i.e., a patient-reported outcome measure, or PROM), a healthcare professional (i.e., a clinician-reported outcome measure, or ClinROM), or both. This decision will not be considered binding for the final CMS owing to the uncertainty at this stage around the availability and quality of candidate measures. The decision instead will be used during Phase 4, to help inform the selection of instruments for the CMS.

### Phase 2. Identifying potential instruments and evaluating their measurement properties

Phase 2 will be conducted in three stages: (2.1) a systematic review to assess the quality of existing measurement instruments used in DCM; (2.2) a gap analysis of elements, to identify those for which a measurement tool of sufficient quality within DCM does not exist; and (2.3) targeted scoping reviews of these gap elements, to identify potentially relevant instruments used outside of DCM.

Phases 2.1 and 2.2 have been completed. Phase 2.1 will be published separately; thus, only a summary is provided here. Phase 2.2 and its results are included here.

#### 2.1 Systematic review of existing measurement instruments

A systematic review was used to evaluate the quality of a predefined list of existing measurement instruments, identified from three previous scoping reviews (13, 33, 34). The term ‘measurement instrument’ was used to refer to how the element was being measured (i.e., the instrument used to assess the outcome) and could refer to a single question, a questionnaire, or other tools (35, 36), including PROMs and ClinROMs.

The search was performed in EMBASE and MEDLINE from inception until 4 August 2020 to identify original research assessing the measurement properties of instruments used in clinical research of DCM. The search string was built using the relevant DCM search filter (37, 38) and the COSMIN filter for studies evaluating measurement properties (39). Abstracts were screened by four reviewers against a set of pre-defined criteria (**Supplementary Table 1**). Only primary clinical research studies evaluating one or more measurement properties were included.

All data were collected, processed, and analysed in accordance with the COSMIN manual for systematic reviews of PROMs. This involved collecting results across 10 measurement properties: content validity, structural validity, internal consistency, cross-cultural validity/measurement invariance, reliability, measurement error, criterion validity, hypotheses testing for construct validity, responsiveness, and clinically important differences. Results were rated as ‘sufficient’, ‘indeterminate’, or ‘insufficient’ and overall methodological quality scores were scored as ‘very good’, ‘adequate’, ‘doubtful’, ‘inadequate’, or ‘not applicable’, as described in the manual. Results were then qualitatively summarised and an overall rating of the quality of the studies was made using a modified Grading of Recommendations Assessment, Development, and Evaluation (mGRADE) approach, as described in the manual. Recommendations were formulated based on all evidence, a list of interpretable instruments was collated, and findings were subsequently reported as a narrative synthesis (40).

#### 2.2 Gap analysis

Whilst the review identified clinically interpretable instruments that were common to DCM research and could be used to measure outcomes in the COS, there were: (a) several elements for which no existing tool was appropriate and (b) several instruments for which the evidence base was deemed inadequate (23).

To identify candidate tools for these gaps, we looked for appropriate tools outside of the field of DCM. Before conducting scoping reviews for each gap *de novo*, a pragmatic MEDLINE search was performed to assert if such reviews already existed. Outcomes within the domain of pain were excluded as it was felt the resources and recommendations aggregated by the Initiative on Methods, Measurement and Pain Assessment in Clinical Trials (IMMPACT) were sufficient (41). Search strings were formed, comprising the core outcome, synonyms of ‘psychometric’ and ‘neuroscience’ (37, 39), and were limited to the last five years to ensure relevance. The search was restricted to neuroscience, as it was anticipated this would most likely identify tools with appropriate content validity. Abstracts were screened by one reviewer against the same criteria from the review (**Supplementary Table 1**). Results from this gap analysis are aggregated in **Table 2**. Notably, no systematic reviews were identified, but a published protocol with respect to fatigue was (42) obtained via personal communication.

**Table 1.**
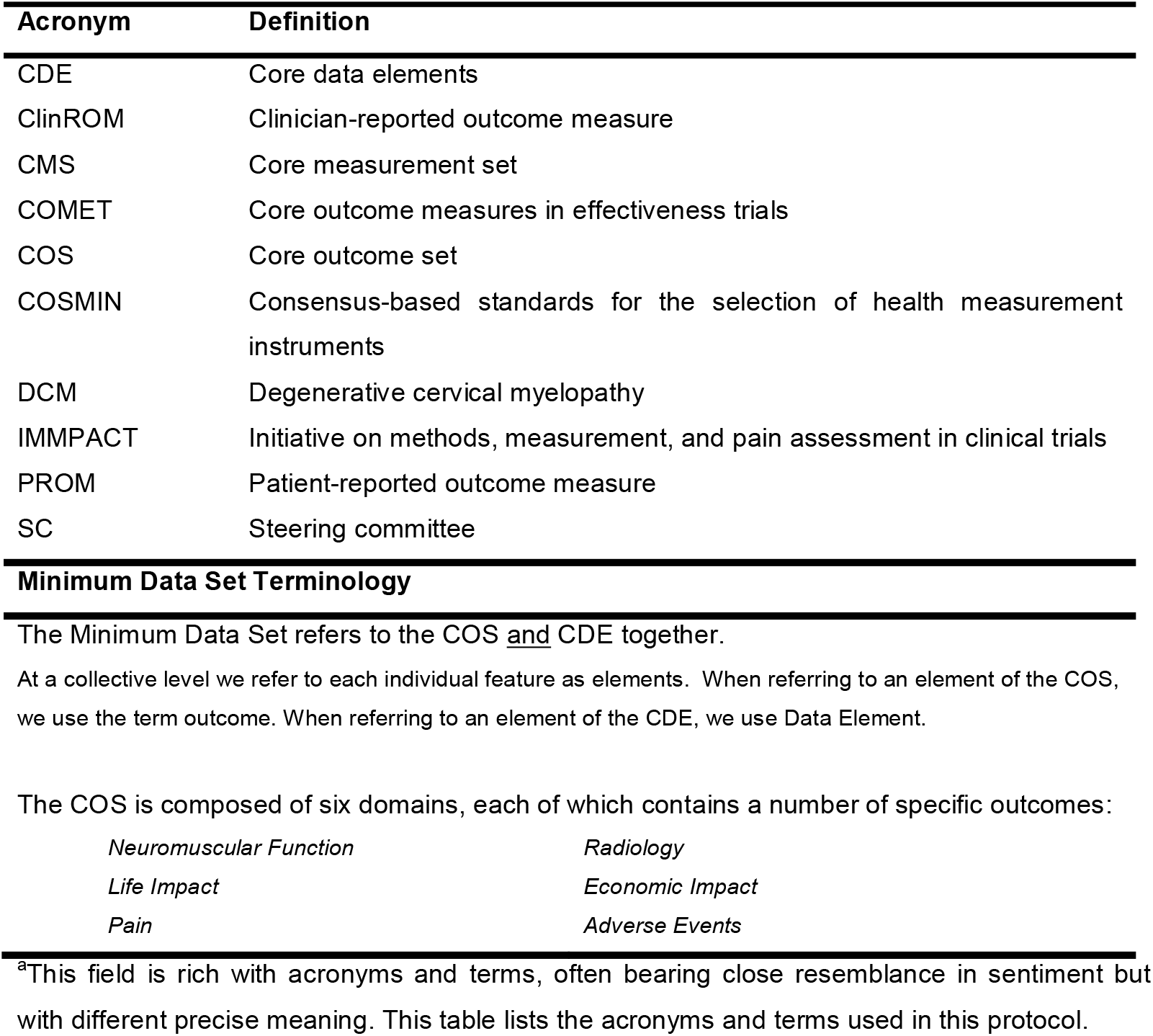
RECODE-DCM Definitions and Terminology.

**Table 2.**
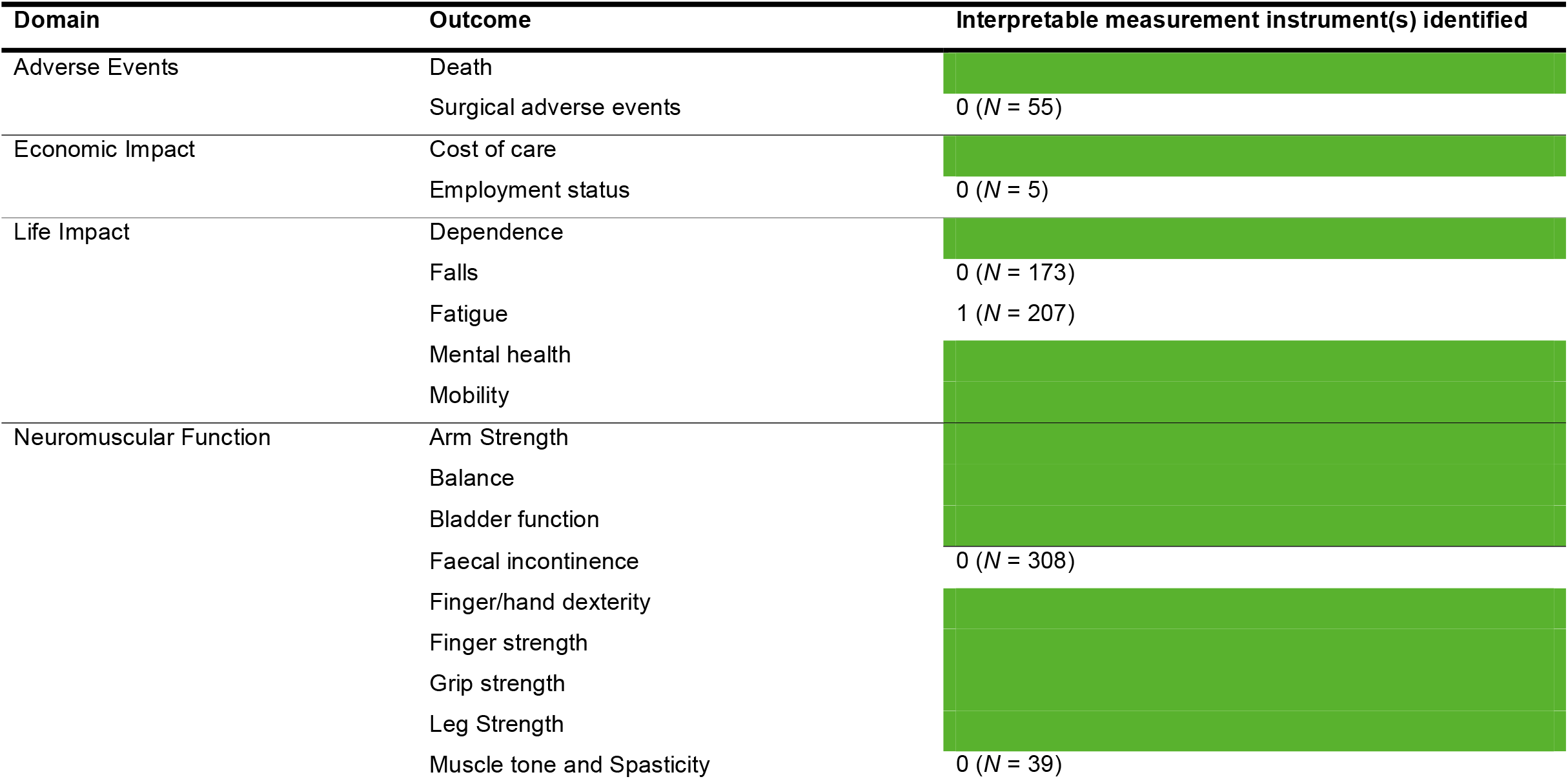
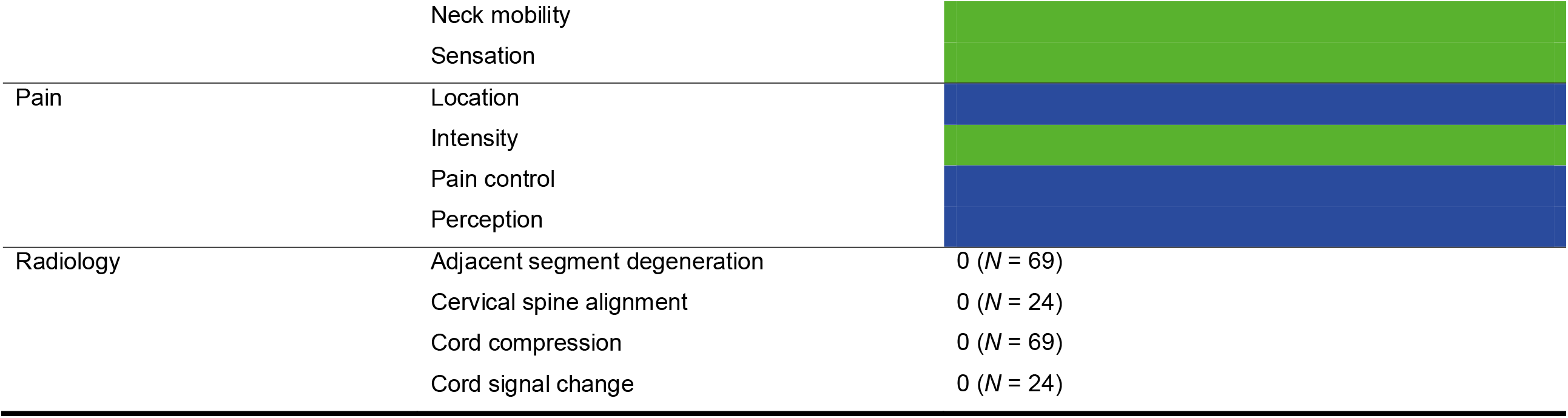
Gap analysis. Elements with at least one interpretable instrument (see Phase 2.1) are shaded green and will be published separately. Targeted searches of MEDLINE were performed for the remaining elements (i.e., ‘gaps’, unshaded, see Phase 2.2). For gaps within the domain of pain (shaded blue), the resources aggregated by IMMPACT were deemed sufficient (43). The number of articles (*N*) screened is indicated for each gap. Notably, only one suitable resource was identified for ‘fatigue’.

#### 2.3 Targeted scoping reviews

For those remaining outcomes without potential instruments, focused scoping reviews will be conducted using MEDLINE and EMBASE. These reviews will be conducted in two stages and will aim: (a) to identify instruments used in a related target population; and (b) to evaluate the methodological quality of those identified tools.

Searches will be conducted in disease populations likely to measure the same construct. For example, ‘faecal incontinence’ could be a symptom of many diseases. However, since this symptom is also measured in other spinal disorders with neurological injury (e.g., traumatic spinal cord injury and cauda-equina syndrome), these disorders would be considered appropriate populations. These will be defined with input from stakeholders a priori.

As a scoping exercise, this initial search will focus on reviews to develop lists of measurement instruments. These identified tools will then be specifically combined with the COSMIN filter (39) and the chosen target population, to aggregate studies evaluating their measurement properties. In addition, these tools will be searched in the COSMIN database.

### Phase 3. Evidence on timing of assessment

The timing of the assessment is an additional source of variation with respect to aggregating outcomes. Due to the current uncertainty around the natural history of DCM (recognised as a critical research priority) (44), analysing timing of assessment will not be possible for studies considering non-operative management. However, for DCM managed operatively, the recovery profile is more stereotyped and is felt more amenable to the standardisation of measurement time points.

To help inform this recommendation, an evaluation of the AO Spine Cervical Spondylotic Myelopathy (CSM) North America and International datasets will be conducted (45, 46). These are two high-quality observational studies of patients undergoing surgery for DCM, followed up at three, six, twelve, and twenty-four months after surgery. These incorporate the most frequently used follow-up timepoints from DCM research (13). Recovery trajectories will be modelled over time, including the proportion of patients achieving maximal recovery at each follow up point and the percentage change from last follow up. The significant contextual factors that may influence this (e.g., age or comorbidities) will also be explored. These findings will be shared during Phase 4.

### Phase 4. Consensus recommendations

#### 4.1 Formation of an expert consensus panel

A multi-disciplinary panel of experts will be formed to finalise the CMS through consensus. These experts will be identified using purposive sampling to include people with lived experience; professionals from key clinical disciplines commonly involved in DCM care (i.e., spinal surgery, neurology, rehabilitation medicine, physiotherapy, and primary care) (12, 47); professionals with clinical trials experience, particularly with respect to measuring each of the six domains (i.e., adverse events, economic impact, life impact, neuromuscular function, pain, and radiology); and professionals with experience in trial statistics. At least half of all participants will be external to the SC; at least one in five participants will have lived experience; and no more than half of all participants will be spinal surgeons. It is also intended to have a 1:1 ratio of women to men. All panellists must declare any conflicts of interest, and be approved by the SC.

#### 4.2 Pre-meeting short-listing

Panellists will be provided with a summary containing the identified measurement instruments considered of sufficient quality for each element, including their evidence base, and the original SC decision concerning the preferred reporting method (i.e., PROM or ClinROM). Each panellist will be asked to submit three preferred measurement tools in advance of the meeting, with a justification for each, and to optionally suggest unrepresented tools.

#### 4.3 Face-to-face consensus meeting

A consensus meeting of the panel will then be convened. The aims will be to: (a) select the preferred measurement instruments, (b) define how they should be reported, and (c) outline when they should be reported in surgically treated DCM cohorts. The management group will prepare documentation for each outcome, comprising those tools shortlisted by the panel during Phase 4.2, together with their evidence. Each outcome will be discussed in turn with a majority decision considered consensus agreement. The consensus meeting will be overseen by an independent facilitator and follow a nominal group technique. Moderated discussion and re-voting will be undertaken as necessary until consensus is achieved for all components of the COS and CDE. Consensus will be defined as >70% agreement.

### Phase 5. Implementation

The dissemination of the CMS will be incorporated into the active knowledge translation proposal for the entire AO Spine RECODE-DCM initiative. This includes scientific publication, conference presentations, podcasts, identifying AO Spine RECODE-DCM ambassadors, and engaging with relevant journals and funders. This process will be subject to periodic review to ensure strategies are effective and adaptive.

This will include a survey of the RECODE-DCM community, designed to share the CMS and ascertain barriers to implementation. This information will be used to inform overall strategy.

The AO Spinal Cord Injury Knowledge Forum, an international and multidisciplinary group of professionals working in this field, will review the relevance of the CMS at 4 years from release, to consider whether an update is required.

## DISCUSSION

This protocol outlines the process for developing a CMS for DCM, based on the CDE and COS already defined by AO Spine RECODE-DCM. Whilst some pragmatic steps have been taken, this process remains faithful to consensus methodology and CMS precedent (23, 25-32, 35) and, ultimately, remains robust.

### The CMS will focus on measurement instruments currently in usage

From the outset, it was decided that the CMS would principally focus on existing instruments currently in use. Although the development of better assessment tools is a top 10 research priority (24), the strategy to use existing tools was preferred for several reasons. First, the aim of this project was to develop a CMS that could be immediately implemented in clinical practice and research studies. The development of new tools remains a work in progress, including microstructural MRI, gait laboratory analysis, and clinical assessments (24, 48, 49). Whilst it seems inevitable that these measurement instruments will change DCM assessment, there remain important methodological uncertainties, practical challenges, and technological requirements that pose potential barriers to adoption.

Widespread adoption is necessary for a minimum data set to improve research efficiency. Unless individual DCM researchers have unified data collection, the comparison of findings across studies will remain limited (50). Changing practice, however, is challenging, particularly when a concept is unfamiliar or questioned (51-53). It is therefore important to recognise that CMSs can be updated (54) and that individual studies can incorporate additional tools at their discretion. Furthermore, the inclusion of emerging technology should only be included in future CMS iterations when their selection is undisputable.

For DCM, an equally important but more achievable priority is to ensure that the intended breadth of outcomes is being measured. As highlighted in Phase 2.2, previous studies may have underrepresented the disease. (13, 18). This holds significant implications for interpreting the literature. A recent example is the results of the CSM-Protect study, a randomised controlled trial comparing riluzole as an adjuvant to surgery to surgery alone (55). While there were no differences between treatment groups with respect to the primary endpoint (i.e., neuromuscular function), there were indications of meaningful benefit amongst secondary outcomes (e.g., complications such as C5 nerve palsy, and pain).

As a nascent research field with a paucity of high-quality prospective studies (9, 56), ensuring that current research is comparable to these benchmarks will be important for their generalisation and implementation in the short-term (17). This will require existing measurement instruments to be represented.

### The CMS will be selected using nominal group techniques

Several methods exist to achieve meaningful consensus (57, 58). Ultimately, these methods aim to ensure that all relevant perspectives are captured and appropriately represented in the decisions taken (59). Consensus processes are increasingly approached by combining literature evidence, serial surveys, and a final consensus meeting—a modified Delphi (57, 60, 61). This approach was effectively used during our previous three consensus processes (i.e., for the index term, CDE, and COS).

The diverse perspectives from different stakeholder groups was imperative in determining ‘what’ to measure, identifying previously unprioritised outcomes (62) and developing a global multi-stakeholder community focused on DCM (63). Arguably, ‘how’ to measure these outcomes will require further focused perspectives on clinical assessment and trials. When conducting our international Delphi processes, engaging under-represented stakeholders was challenging (12, 64, 65). At the outset, we aimed to capture perspectives of people with lived experiences, surgeons, and other healthcare professionals in a 2:1:1 ratio (12). However, this could not be achieved, and engaging spinal surgeons—who most frequently treat, research, and specialise in DCM—was much easier (65). Given that the CDE and COS have been defined, and that the decision on how to measure them is likely to benefit from specific expertise, a purposively selected group using a nominal group technique was favoured for the CMS. It is also hypothesised that the step of sharing the results of the CMS with the wider DCM research community will facilitate dissemination and improve face validity.

## CONCLUSIONS

This protocol describes the formation of the first CMS for DCM, which will focus on instruments in current use. This aims to facilitate the standardised and comprehensive measurement of DCM and will create a venue for the development and introduction of novel measures. We anticipate that this process will greatly facilitate knowledge generation and knowledge translation in DCM by enabling clinicians and researchers to ‘speak a common language’ with regard to outcome instruments.

## Data Availability

All data produced in the present work are contained in the manuscript.

## ACKNOWLEDGEMENTS

We thank the ongoing support of our wider stakeholders, including AO Spine RECODE DCM Community, and partners, including Myelopathy.org (DCM Charity; www.myelopathy.org). Further information about the initiative, and opportunities to get involved can be found at www.aospine.org/recode

## CONTRIBUTORS

BMD was responsible for conceiving the article. AM contributed to the study design. BMD and AYT wrote the protocol and manuscript and contributed equally to this paper. BK, MGF, MRNK, and IS facilitated international collaboration. BMD, AYT, ODM, KSL, DK, JCF, MGF, JH, CMZ, RRP, JM, ES, AC, VRM, BA, TFB, LT, RC, JDG, SKR, IS, SW, AGKM, MRNK provided critical appraisal of the manuscript. All authors critically revised and approved the manuscript.

## ETHICS APPROVAL

Ethical approval for this project was obtained from the University of Cambridge (Ethical approval number: HBREC2019.14)

## DATA AVAILABILITY STATEMENT

All data produced in the present work are contained in the manuscript.

## FUNDING

This work was supported by AO Spine through the AO Spine Knowledge Forum Spinal Cord Injury, a focused group of international Spinal Cord Injury experts. AO Spine is a clinical division of the AO Foundation, which is an independent, medically guided not-for-profit organisation. Study support was provided directly through the AO Spine Research Department.

## COMPETING INTERESTS

None declared.

## Notes

### Competing Interest Statement

The authors have declared no competing interest.

