## Supplementary Table 1 for "Development of a core measurement set for research in degenerative cervical myelopathy: a study protocol (AO Spine RECODE-DCM CMS)"

### SUPPLEMENTARY INFORMATION

**Supplementary Table 1. Inclusion and exclusion criteria for the systematic review.**

| Inclusion | Exclusion |
|---|---|
| Publication type | |
| • Article written in English<br>• Primary clinical research articles | • Article not written in English<br>• Conference abstracts or posters<br>• Editorials, commentaries, opinion papers or letters<br>• Book chapters or theses |
| Study type | |
| • Study includes primary clinical data | • Study uses only secondary data<br>• Case reports<br>• Narrative reviews<br>• Systematic reviews<br>• Meta-analyses |
| Populations | |
| • Human studies | • Non-human studies |
| Indications | |
| • Exclusively DCM (CSM, ossification of the posterior longitudinal ligament, cervical stenosis, spondylosis, spinal cord compression, cervical myelopathy) | • Populations with DCM and at least one other condition (e.g., radiculopathy) |
| Comparator | |
| • At least one assessment tool from (11, 30, 31) | |
| Outcomes | |
| • At least one psychometric property<br>• At least one MCID or SCB | |
